# Donation strain engraftment demonstrates feasibility of vaginal microbiota transplantation to prevent recurrent bacterial vaginosis

**DOI:** 10.1101/2025.08.27.25334544

**Authors:** Fatima A. Hussain, Agnes Bergerat, Julia Kelly, Briah Cooley Demidkina, Daniel Worrall, Jiawu Xu, Aditi Kannan, Tess Brunner, Natalie Culler, Miles Goldenberg, Wafae El Arar, Ankita V. Katukota, Meena Murthy, Joseph Elsherbini, Salina Hussain, Mary Dong, Douglas S. Kwon, Caroline M. Mitchell

## Abstract

Although bacterial vaginosis (BV) affects 30% of women worldwide and is associated with adverse health outcomes, current standard-of-care antibiotics fail in over half of cases and treatments have not improved in over 40 years. Probiotics have been proposed as alternative treatments, but fail to restore an optimal lactobacilli-dominated microbiome in the vast majority of patients. Here, we present findings from a pilot clinical trial demonstrating the successful engraftment of vaginal microbiota transplantations (VMTs) after antibiotic treatment in individuals with recurrent BV. Following an investigational donation protocol under an FDA IND, we treated eight recipients with material from a single donor. Using 16S rRNA gene amplicon sequencing we show that VMT results in a shift toward an optimal, *Lactobacillus crispatus*-dominated microbial community in three out of four VMT recipients at one month post-transplant. In two successful transplantations, this shift lasted at least six months post-VMT. In contrast, no placebo recipients exhibited *L. crispatus* dominance. Bacterial culturing and whole genome sequencing combined with metagenomic sequencing from donations and recipient longitudinal samples revealed colonization by donor-derived strains of *L. crispatus* in VMT recipients. Additionally, we observed no increase in genital inflammatory markers or changes in endocervical immune cell proportions when comparing treatment to placebo, indicating transplant safety. Together, these findings support the hypothesis that transferring the entire vaginal microbiota can lead to a more complete restoration of the vaginal ecosystem compared to single strain probiotics and lay the foundation for designing novel microbial therapies for BV.

*Vaginal microbiota transplantations lead to stable L. crispatus engraftment in the microbiomes of certain patients with recurrent bacterial vaginosis.*

## Introduction

Bacterial vaginosis (BV) is a clinical syndrome which impacts over a quarter of women worldwide and is associated with adverse reproductive health outcomes such as preterm birth, acquisition of HIV, and progression of HPV-related cervical dysplasia (*1–4*). BV symptoms include vaginal discharge, odor, and irritation which significantly reduce quality of life, sexual health, and self-esteem. The antibiotics metronidazole and clindamycin are the current standard therapies for BV, but unfortunately over 60% of people experience BV recurrence within six months of antibiotic use (*5–7*). Despite high rates of recurrence, there have not been new classes of drugs for the treatment of BV in over 40 years (*8, 9*).

The vaginal microbiomes of people with BV typically have low abundances of beneficial *Lactobacillus* spp., and high abundances of multiple species of diverse anaerobic bacteria. One possible reason for high BV recurrence is the lack of vaginal colonization post antibiotic treatment by protective species such as *Lactobacillus crispatus*, a species associated with beneficial health outcomes in populations worldwide (*10*). After successful antibiotic treatment, a diverse BV-associated microbiome is often shifted to an ecosystem dominated by *L. iners,* a less protective species of *Lactobacillus*, which is frequently associated with transition back to a diverse BV-associated microbiome. As such, there is great interest in supplementing antibiotic treatment with a probiotic or a live biotherapeutic product (LBP) containing *L. crispatus*.

An ideal product would provide durable benefit after short term use through the sustainable colonization of health-associated bacteria. In a clinical trial of a single strain *L. crispatus* vaginal LBP (Lactin-V), people receiving the LBP for 12 weeks after antibiotic treatment had a significantly lower rate of BV recurrence than those receiving placebo (*6*). However, recurrence 6 months after antibiotic treatment in the LBP arm was 39%, and only 44% of participants retained colonization with the LBP strain. No over-the-counter probiotics for vaginal health have been studied as rigorously as Lactin-V, and few of those studied contain species of *Lactobacillus* commonly found in the vagina (*11*). Finally, few other studies have assessed the long term impact of these products after cessation of use.

Vaginal microbiota transplantation (VMT) has been proposed as a novel therapy to effectively and durably establish colonization with optimal vaginal species of the genus *Lactobacillus* (*12, 13*). The precedent of fecal microbiome transplantation (FMT) to effectively treat antibiotic refractory infections in the gut demonstrates that transfer of a whole microbial community can be more effective than probiotics. A case series of VMT in people with highly recurrent BV showed *L. crispatus* colonization and dominance in four out of five participants after one to three doses, which was stable for 5-21 months (*13*). In a population with a history of multiple BV recurrences, this was a remarkable achievement. However, the study lacked a placebo comparison and used 16S rRNA gene sequencing alone to profile bacteria, which precluded confirmation of colonization by specific donor-derived *Lactobacillus* strains.

Here, we present a pilot Phase 1 randomized trial of VMT in people with recurrent BV. We demonstrate significantly greater *L. crispatus* dominance after VMT vs. saline placebo one month post-transplant, and stable engraftment with donor-derived *Lactobacillus* strains lasting at least 6 months in half of VMT recipients. By investigating microbial and host factors across all study participants, we generate testable hypotheses to differentiate VMT responders from non-responders, and propose design principles for novel microbial therapies for BV.

## Results

### Study design and comprehensive screening of VMT donor and donations

We enrolled and randomized eight participants with a history of recurrent BV, defined as two episodes in the past six months, or three episodes in the past year, and a Nugent score greater than four at enrollment (timepoint/visit, V01) (Table 1, Figure 1A). At their baseline visit (V02), all participants reported at least one mild symptom, typically vaginal odor and/or discharge (Supplemental Data S1). All participants then received 500mg oral metronidazole twice daily for seven days as the standard-of-care treatment, which was timed as closely with the duration of menses as possible. Within 24-72 hours of the final dose of antibiotics, participants returned to receive the first VMT (V03), and 36-72 hours later received a second VMT dose (V04). Participants then returned one week (V05), two weeks (V06), and four weeks post-transplant (one month, V07), as well as two months (V08), four months (V09), and six months post-transplant (V10) for follow up visits (Figure 1A). At each follow up visit, participants completed a health and behavior questionnaire to evaluate symptoms and adverse events as well as provided vaginal swabs and/or menstrual disc samples for microbiome analyses (Methods). Bacterial 16S rRNA gene amplicon sequencing was performed on DNA extracted from one swab taken at the beginning of each visit, and used to evaluate the primary outcome of the study—*Lactobacillus* dominance (>50% relative abundance) at one month post VMT (V07). The extracted DNA from each sample was also used for metagenomic sequencing to test for strain-level engraftment as well as qPCR to measure absolute abundances of different bacterial species. Menstrual disc samples collected at visits V02, V03, V05, V07, and V09 were used to measure inflammatory cytokines, and cytobrush samples collected at visits V02 and V07 were used to measure immune cells directly using flow cytometry.

**Table 1.** Recipient demographics and baseline characteristics.

|  | **VMT** | **Placebo** |
| --- | --- | --- |
| **Age Median (Range)** | 30 (26-33) | 39 (32-43) |
| **Self-identified Race** |  |  |
| Black | 3 (75%) | 1 (25%) |
| White | 1 (25%) | 3 (75%) |
| **Hispanic/Latinx** | 1 (25%) | 0 (0%) |
| **Gender of sexual partners** |  |  |
| Male | 2 (50%) | 2 (50%) |
| Both male and female | 1 (25%) | 1 (25%) |
| No current partner | 1 (25%) | 1 (25%) |
| **Sexual partners in past month Median (Range)** | 1 (0-2) | 1 (0-1) |
| **Contraceptive use** |  |  |
| Abstinence | 0 (0%) | 1 (25%) |
| Condoms | 1 (25%) | 2 (50%) |
| Copper IUD | 0 (0%) | 1 (25%) |
| Levonorgestrel IUD | 2 (50%) | 0 (0%) |
| Oral contraceptive pills | 1 (25%) | 0 (0%) |
| **Nugent at enrollment Median (range)** | 7 (4-8) | 6 (5-7) |

**Fig. 1.**
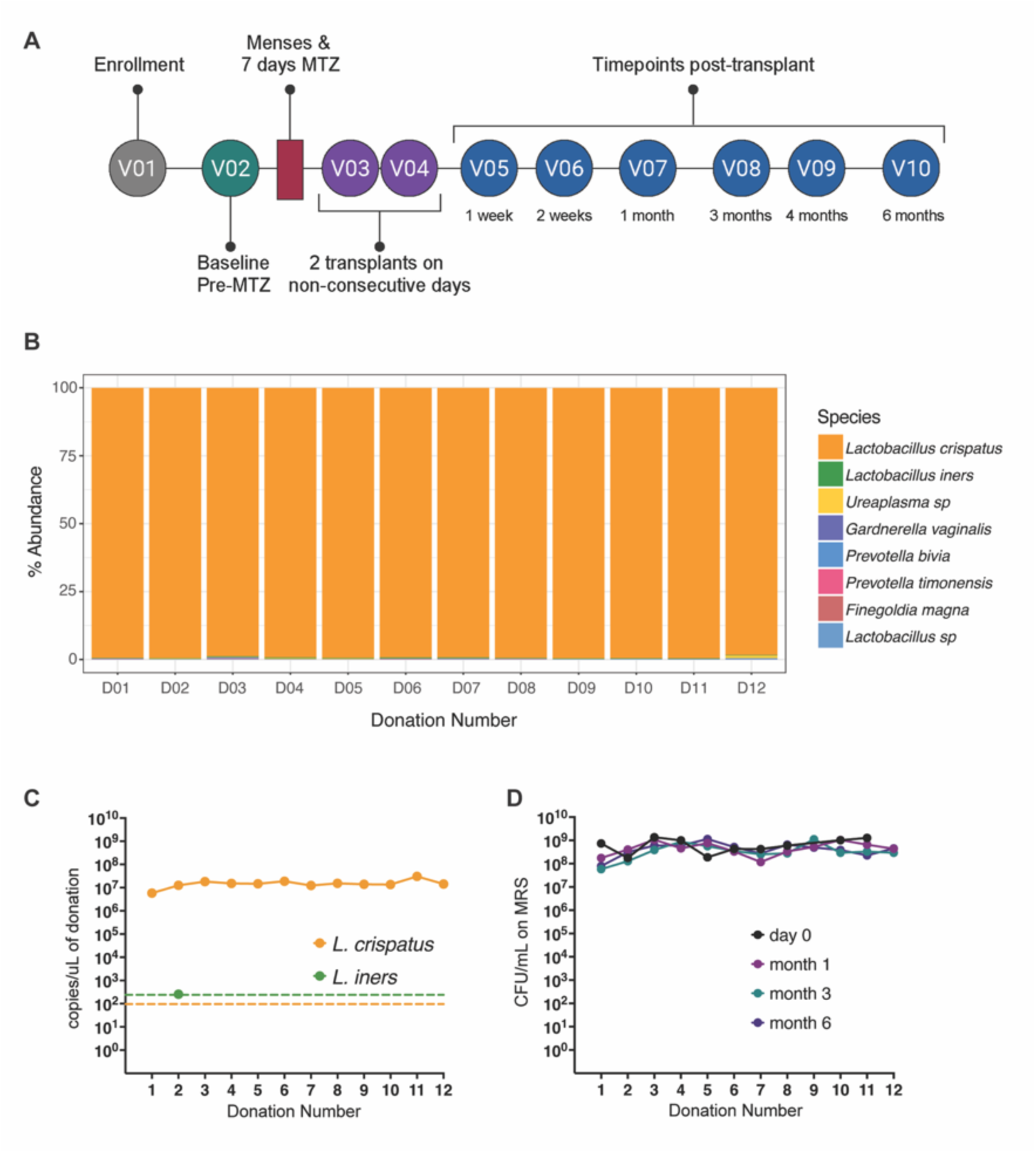
VMT trial design and donation screening. (A) Study schematic depicting all visit timepoints. (B) Bacterial profile of each VMT donation characterized with16S rRNA amplicon sequencing. (C) Absolute abundance of *L. crispatus* and *L. iners* using qPCR, limit of detection depicted as dashed lines. (D) Stability of donation material in -80°C storage over 6 months, measured as colony forming units grown on MRS agar.

A single donor provided all the donations used in this pilot study. The donor is a healthy, reproductive age woman who had never had BV and tested negative for a comprehensive panel of infections in our donation protocol, conducted under an FDA IND (*14*). The donor collected vaginal fluid using a disposable menstrual disc that was inserted and worn overnight, which was then processed and tested as previously described (*14*) and stored at -80°C without added cryoprotectants until cleared for use. The donor’s microbiome throughout the time of sample collection was stably, and nearly completely, dominated by *L. crispatus* – as measured by 16S rRNA gene amplicon sequencing for relative abundance (Figure 1B). Congruently, the absolute abundance of *L. crispatus*, measured by qPCR, was at least four orders of magnitude higher than that of *L. iners* across all donations (Figure 1C). Donation aliquots were tested for stability of bacterial viability throughout the storage period (Figure 1D). After completing all safety testing, and microbiome analysis, each individual donation was used to create a single dose with a volume of at least 700uL. A single VMT dose was cleared for use only when the donor completed the final post-donation testing, and the dose passed all quality checks. The range of total colony forming units (CFUs) delivered in a single dose of VMT was 2.8 x 10^8^-6.6 x 10^8^ CFUs, and donations ranged in volume from 1.0-2.1 mL and in weight from 0.8-2.0 g (Figure S1).

### Recipient transplantation safety and microbiome and immune dynamics

There were no serious adverse events in the pilot study. There were two Grade 2 genitourinary adverse events in the placebo arm, and zero in the VMT arm (Table S1). Prior to antibiotic treatment, only 1/4 (25%) of placebo recipients (participant, P04) had *L. crispatus* detected via 16S rRNA gene amplicon sequencing at greater than 1% relative abundance (1.2% at V01 and 18.2% at V02) (Figure S2). At baseline (V02), 2/4 (50%) people in each group had *L. iners* dominance (>50% relative abundance). Our primary outcome of *Lactobacillus* dominance at four weeks post-cessation, measured via 16S rRNA gene amplicon sequencing, was achieved in 4/4 (100%) VMT recipients and 1/4 (25%) placebo recipients, while our secondary outcome of *L. crispatus* dominance at four weeks, was achieved in 3/4 (75%) VMT recipients and 0/4 (0%) placebo recipients. Notably, all four VMT recipients established *L. crispatus* dominance in at least one sample in the four weeks after VMT, and *L. crispatus* dominance persisted up to at least six months in 2/4 (50%) VMT recipients. In the placebo group no participants ever attained *L. crispatus* dominance.

In addition to 16S rRNA amplicon sequencing, we performed taxonomic profiling using metagenomic sequencing data which provided additional genomic resolution in certain cases. For example, 16S sequencing is unable to distinguish *L. crispatus* from *L. acidophilus*, which was disambiguated by metagenomic profiling in a participant in the placebo arm (P03) who surprisingly had a high abundance of *L. acidophilus*, which is not commonly found at this abundance in the vaginal microbiome (Figure 2). Due to the higher genomic resolution, metagenomic sequencing data was also used to measure the increase in relative abundance of *L. crispatus* to track this specific microbiome shift of interest (Figure S3). Finally, absolute abundances of each species, measured using species-specific qPCR, agreed with relative abundance of each species of interest (Figure S4).

**Fig. 2.**
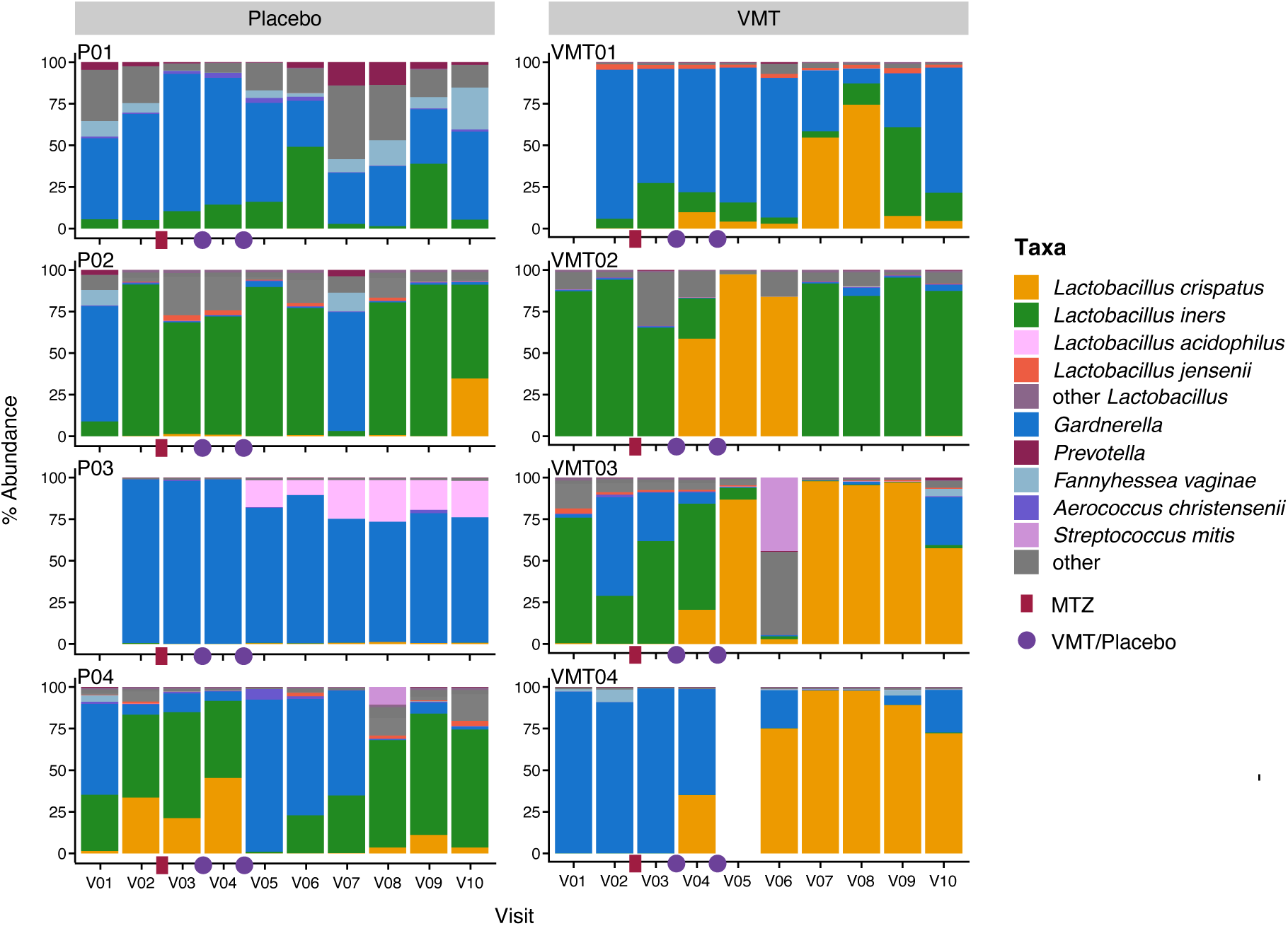
Microbiome shifts in placebo versus VMT recipients. Vaginal microbiome relative abundance in recipients treated with placebo (left) or VMT (right). Stacked abundance bar graphs display the community composition of each sample (taxonomic assignments from VIRGO2 (*18*)). Metronidazole administration depicted with red bar. Dosage of VMT or saline placebo indicated by purple circles.

In this pilot study, we observed no obvious relationships between either behavioral metadata, or microbiome community (pre- or post-metronidazole) and VMT success. For example, the one VMT recipient who did not retain *L. crispatus* at one month (VMT02) did not report more or different sexual exposures than the other three VMT recipients, nor did she have a more diverse pre-treatment microbial community, or more severe symptoms. She had the lowest frequency of detection of classic BV-associated species such as *G. vaginalis* and *F. vaginae*, but had the highest relative abundance of *L. iners* prior to metronidazole treatment. Her absolute abundance of *L. iners* was 9.97×10^9^ copies/swab by qPCR, about 3-fold higher than that of the next closest recipient (VMT03). The second participant who lost *L. crispatus* dominance before 6 months retained colonization with donor-derived *L. crispatus* isolates. All three people with *L. crispatus* dominance at one month had concurrent detection of *G. vaginalis*.

Because secretor status, which is indicative of whether a person secretes ABO blood antigens into their mucus, has been correlated with composition of the vaginal microbiome, we also tested if secretor status of the recipients correlated with VMT success. We evaluated secretor status by using a specific restriction enzyme to cut the PCR-amplified Fut2 gene and observing predicted banding patterns based on expected mutations (Methods). The donor, along with all recipients of VMT and placebo except for VMT02, have a secretor phenotype (Supplemental Data S1).

When comparing vaginal fluid cytokines after VMT vs. after placebo we did not observe any significant differences, either in absolute concentrations (Figure S4) or in shifts from baseline concentrations (Figure S5). These observations were corroborated with the fact that we also did not see a significant change in endocervical immune cell populations after VMT compared to placebo (Figure S6). This is reassuring from a safety standpoint, in that VMT did not appear to cause vaginal mucosal inflammation.

### Donor strain engraftment in VMT recipients demonstrates transplant success

To test for *L. crispatus* engraftment from the donations into the VMT recipients, we used comparative genomics of donor and recipient strains. We isolated 10 colonies on *Lactobacillus*-selective MRS media from each donation, as well as 10 colonies from each recipient sample where *L. crispatus* showed >2% relative abundance by 16S rRNA sequencing. Isolates were sequenced using both Illumina short read sequencing and Oxford Nanopore long read sequencing to allow for hybrid, high quality genome assembly. The genomes were then compared between the donor and the pilot participants by constructing a phylogenetic tree based on core genome alignment (Figure 3A). Given that the genomes of the VMT donor strains and VMT recipient strains are indistinguishable on the tree and cluster separately from the genomes of the strains isolated from placebo recipients and other reference strains, we can infer that the *L. crispatus* isolates from VMT recipients were indeed the same as those isolated from the donor (Figure 3A). Of note, the two VMT recipients who did not have *L. crispatus* dominance at 6 months both colonized with the donor-derived isolates for at least two visits, and one retained colonization with donor-derived isolates at 6 months despite *L. crispatus* not being dominant (VMT01).

**Fig. 3.**
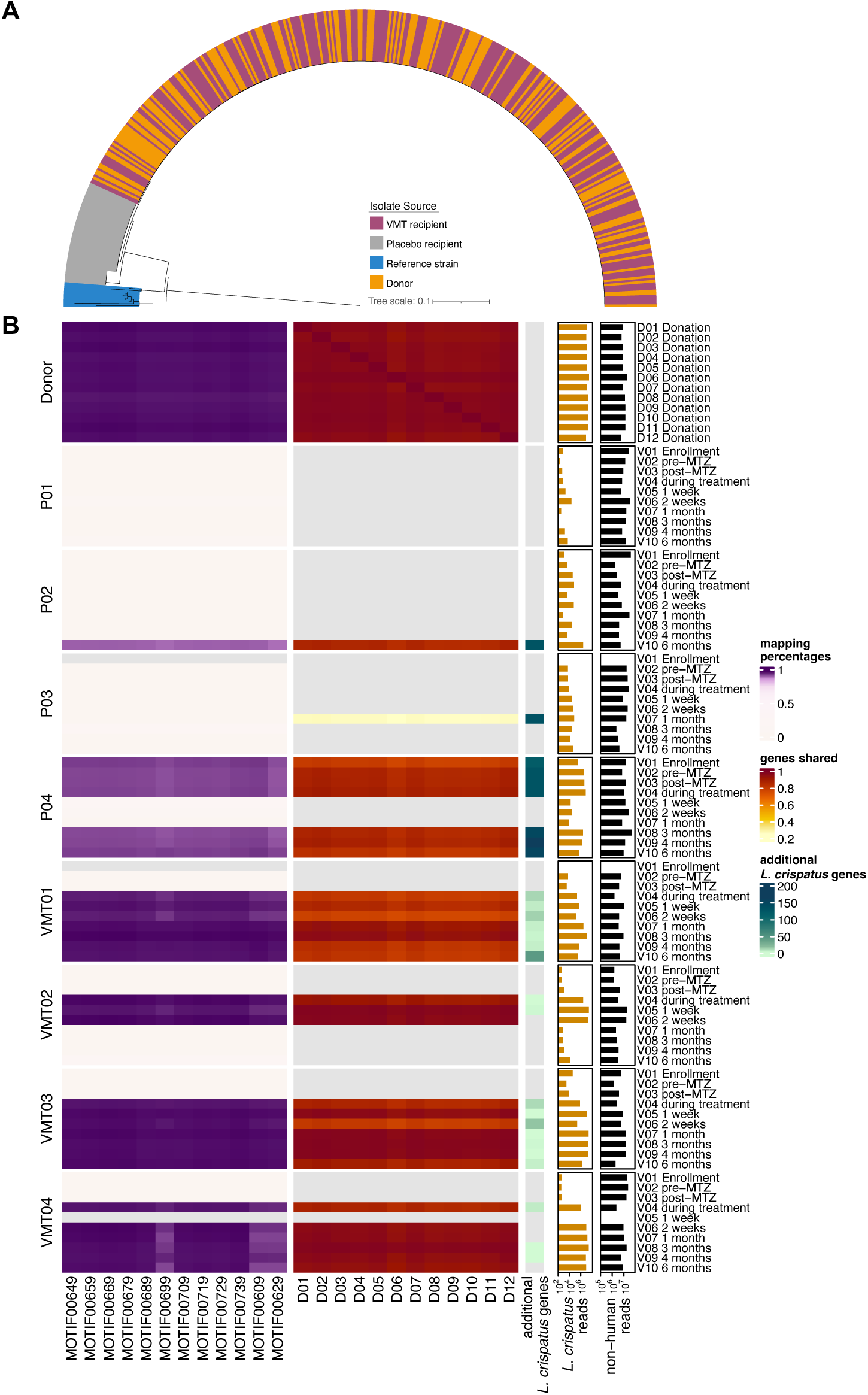
Strain-level analysis of genomes and metagenomes identifies donor strain engraftment in VMT recipients. (A) Whole genome tree of *L. crispatus* strains isolated from VMT Donor (orange), VMT recipients (magenta), and Placebo recipients (grey), with reference strains as the outgroup (blue). (B) Metagenomic analysis of donations and recipient samples confirm engraftment of VMT. Each row represents a donation or recipient sample metagenome. From right to left: Black bar graphs display total number of human-filtered reads in each sample. Orange bar graphs display total number of *L. crispatus*-specific reads in each sample (determined by the VIRGO2 gene catalog). The teal gradient represents the absolute number of *L. crispatus* genes present in each sample which are not present in any of the donation metagenomes (gene is counted as present if coverage is >10% of the median reads per gene). Red gradient columns represent the fraction of *L. crispatus* genes shared between each donation and sample metagenome. Purple gradient columns represent the fraction of each donation isolate genome where sample reads map with >10% of the median coverage.

To confirm that the isolates we analyzed were representative of the sample from which they originated, we compared their genomes to the whole metagenomic sequencing of all donations and recipient samples. We first removed reads from each sample that mapped to a reference human genome and summed the remaining microbiome reads which we expect to cover the bacteria, viruses, and fungi present (Figure 3B, black bars). We then determined the total number of *L. crispatus* reads in each sample by mapping the total host-filtered reads to the non-redundant list of *L. crispatus* genes in the VIRGO2 database (Figure 3B, orange bars). Next, by mapping the metagenomic bacterial reads from each donation to the *L. crispatus* isolate genome from that same sample, as well as to every other donor isolate (Figure 3B, purple heatmap), we were able to confirm that each isolate was present in each corresponding donation, with 95.1-96.0% of the genome having >10% the median coverage at each position. By repeating this analysis for each of the recipient samples, we were able to confirm that the isolates from the donations given to each recipient were present in each VMT recipient sample, with 94.7-96.1% of each genome having > 10% the median coverage at each position. In contrast, only 91.4-92.9% of each genome had > 10% the median coverage at each position for the placebo recipient samples when significant *L. crispatus* was present (>100,000 *L. crispatus* reads), suggesting the presence of an alternative strain not derived from any donations. Note that some proportion of the genome is not well covered with short-read mapping due to repetitive regions.

We then determined how many of the specific *L. crispatus* genes in each donation sample were present in each recipient sample (Figure 3B, red heatmap), and how many additional *L. crispatus* specific genes, not present in the donation samples, were present in the recipient samples of interest (Figure 3B, teal heatmap). As expected, when *L. crispatus* was present in samples from placebo participants, many donation-specific *L. crispatus* genes were absent and many genes that were detected were not from the donations. Samples from VMT recipients with significant *L. crispatus* had very few genes detected that were not from the original donation, and almost all donation genes were present – indicating engraftment of the full genetic potential of the donor *L. crispatus*.

## Discussion

In this eight-person pilot randomized trial of VMT in people with recurrent BV, we demonstrated engraftment of donor *L. crispatus* strains in all VMT recipients, and durable colonization for 6 months in half of VMT recipients. These results provide the most definitive proof to date that after VMT, donor lactobacilli strains establish colonization in recipients. Additionally, we showed no increase in inflammation and no significant adverse events after VMT. This is a clear proof-of-concept that transferring a whole microbial community leads to successful engraftment of vaginal *Lactobacillus* in people with recurrent BV, although the sample size of this study is too small to make definitive conclusions about safety or clinical efficacy. These findings are consistent with safety reported in other studies (*13, 15, 16*). Importantly, continuing to test VMT as an intervention requires robust screening and testing procedures for donors and donations to ensure continued safety for VMT recipients.

The VMT donor in this pilot study had a stable *L. crispatus* dominant vaginal microbial community. If the microbial community was definitive of VMT success, then the VMT should have established the same community in the VMT recipients. However, this is not the case as suggested by this and others’ preliminary work – some VMT recipients do better than others. It is unclear whether this is due to host-microbe incompatibility, an inhospitable recipient microbial community, or something else. Identifying host and environmental factors that determine the composition of the microbial community will shed light on understanding the reasons for VMT success and failure.

This is the first randomized trial of VMT in people with recurrent BV, a population in which it is challenging to achieve cure. The current recommendation for recurrent BV is twice weekly suppressive antibiotic treatment for 3 months, however, 25% of people treated with antibiotics recur during treatment and 50% recur in 3 months after cessation of suppression (*17*), highlighting the inadequacies of antibiotic treatment in this population. Sample size in this pilot is too small to allow definitive conclusions, but it demonstrates feasibility and proof-of-concept. To allow comparison of results across our small number of recipients, we used donations from a single donor, which does not allow comparison of efficacy of *L. crispatus* dominant VMTs between donors, nor how matching of characteristics between donors and recipients impacts VMT success. Most of our recipients did not have clinical BV at the time of initiating antibiotic treatment, so it is unclear whether VMT would have similar effect following antibiotic treatment for acute BV. However, many of our recipients had an *L. iners* dominant bacterial community when they received the VMT, which is common after antibiotic treatment for BV, suggesting these results would be relevant in that clinical scenario.

The success rate in this pilot cohort was similar to that seen in the original case series, which used fresh vaginal fluid instead of fluid that had been frozen and thawed (*13*). Lev-Sagie et al. found 2/5 recipients had a successful transplant with only one donation, 1/5 had a successful transplant with three donations from the same donor, and 1/5 had a successful transplant with three donations where one was from an alternative donor. In their study, 1/5 recipients had a failed transplant. Both studies enrolled people with recurrent bacterial vaginosis and pre-treated recipients with antibiotics. Two recent preprints of randomized trials of VMT in asymptomatic people with a diverse vaginal community, in which no antibiotic pre-treatment was used, did not show a difference in the proportion of people with *Lactobacillus* dominance after treatment. One trial used a single dose of VMT after each of three menstrual cycles, and at 6 months 4/35 (11%) in the VMT arm and 2/11 (18%) in the placebo arm had *Lactobacillus* dominance (p = 0.56) (*15*). The other trial gave three daily doses of VMT, and at 6 months 13/26 (50%) VMT vs 2/8 (25%) placebo recipients had *Lactobacillus* dominance (p = 0.21) (*16*). These data suggest that reduction in the vaginal bacterial load prior to VMT may be important.

BV impacts over a quarter of women worldwide, and despite the dismal long-term cure rates for current antibiotic regimens, treatment recommendations have not substantially changed since the 1980’s. This lack of progress in the field may be due to our lack of biologic understanding of the causes of BV and what determines vaginal *Lactobacillus* colonization. There is no animal model for the human vaginal microbiome, which presents a challenge for studies of causation. Many interventions have been trialed, including Vitamin D supplementation, vaginal lactic acid gel, over the counter oral and vaginal probiotics, vaginal boric acid, vaginal dendrimer gels, vaginal vitamin C, vaginal antiseptics, but none have demonstrated significantly greater efficacy than antibiotics in curing BV. Although the *L. crispatus* live biotherapeutic LACTIN-V significantly reduced BV recurrence compared to antibiotics alone, the LBP isolate did not establish durable colonization in the majority of participants, which may explain the 39% recurrence rate 3 months after cessation of treatment. Our study is too small to make definitive statements about efficacy, but 75% of VMT recipients retained the donor isolates 6 months after receiving just two VMT doses. VMT thus offers not only an opportunity to improve treatment outcomes, but also the opportunity to better understand what factors drive vaginal microbial community composition.

## Data Availability

All data produced in the present study are available upon reasonable request to the authors

## Acknowledgments

**General:** We thank J. Ravel and M. France for providing full access to VIRGO2 database prior to publication, as well as J. Ravel and L. Rutt for guidance on DNA extraction for long read sequencing. We thank all study participants.

## Author contributions

Conceptualization: FH, DK, CM; Methodology: FH, CM, JX, AB, BC, MM, TB; Investigation: FH, AB, JK, BC, DW, JX, AK, NC, MG, TB, WE, AVK, MM, SH; Visualization: FH, JE, AB, CM; Funding acquisition: FH, DK, CM; Project administration: JK, BC, DW, AVK, MM, MD, CM; Supervision: DK, CM; Writing – original draft: FH, DK, CM; Writing – review & editing: all Competing interests: CM has been a consultant for Freya Biosciences, and serves on the scientific advisory board of Ancilia Biosciences and Concerto Biosciences. CM has a financial interest in Ancilia Biosciences, a company developing a new class of Live Biotherapeutics and other bacterial products. Dr. Mitchell’s interests were reviewed and are managed by MGH and Mass General Brigham in accordance with their conflict of interest policies. DK is the CSO of Day Zero Diagnostics.

## Data and materials availability

All data are available upon request. Strains can be made available by requesting an MTA from the corresponding authors.

## Materials and Methods

### Study Design

This pilot, randomized Phase 1a study was conducted at Massachusetts General Hospital between 5/10/22 and 7/12/2023 with the goal of evaluating safety of vaginal microbiome transplantation and establishing proof of concept for the intervention. The study was approved by the Mass General Brigham Institutional Review Board (2019P001543), and all participants signed informed consent. The study was registered on clinicaltrials.gov (NCT04046900), and this safety pilot was pre-specified for unblinding and analysis.

### Participant selection

Participants were recruited through advertisements in clinics, around the hospital, in the local area, on social media, on public transportation, through the electronic medical record, and through referral from physicians within the system. Inclusion criteria were as follows: age 18-55, premenopausal, a history of recurrent bacterial vaginosis (2 episodes in 6 months or 3 episodes in a year), Nugent score at screening of 4-10, and willing to use effective contraception. Exclusion criteria included use of antibiotics, prebiotics or probiotics in the past 30 days (with the exception of metronidazole); allergy to metronidazole; significant medical or gynecologic conditions; BMI > 40; abnormal Pap in the past 12 months; insertion of an IUD in the past 90 days; pregnant, breastfeeding or trying to conceive; HIV infection; or any infection on screening labs that would require treatment with antibiotics.

### Interventions

The vaginal fluid used for transplant was collected, tested, and processed as described previously (*14*). Briefly, healthy, sexually active participants age 18-40 with no prior history of BV and a screening Nugent score 0-3 were tested for bacterial and viral sexually transmitted infections (STI), screened for possible exposure to SARS CoV-2 or Zika virus, had urine toxicology screening for drugs of abuse, and a urine culture and a vaginal fungal culture. Additionally, qPCR for *L. crispatus* and *L. iners* was performed to confirm a ratio of *L. crispatus:L. iners* of >100. Eligible donors then agreed to be sexually abstinent while providing donations. A disposable menstrual disc was placed the night before and then removed upon arrival for their donation visit. Donors could provide up to 20 donations within 45 days of screening labs. Testing for STIs and vaginal yeast was repeated at the final donation visit, and viral serologies were re-tested 30-45 days after the final donation to ensure no acute infection had been missed.

The disposable menstrual disc was centrifuged, the vaginal fluid diluted with 500 µL sterile saline, sheared briefly with a blunt 16g needle, and then aliquoted into analysis aliquots (3 x 25uL, 8 x 10uL). All remaining material was considered the “dose” aliquot, which had to have a minimum volume of 700uL. Sterile saline from the same lot was used to create a placebo dose at the same time. Each donation was tested for high-risk human papillomavirus, seminal fluid, sperm and the *L. crispatus:L. iners* ratio. Doses were stored at - 80°C until use. *Lactobacillus* colony forming units (CFU) were counted from analysis aliquots at 1, 3, 6, and 9 months and/or within 30 days of use of the dose to ensure viability. A drop in CFU count >1 log_10_ was considered a failing result.

### Study procedures

People who signed informed consent and met all eligibility criteria were scheduled for a baseline visit prior to their next menstrual period. At that visit, they were dispensed oral metronidazole and randomized 1:1 to intervention or placebo. Randomization was performed using a computer-generated list, with a block size of four. An unblinded staff member who did not participate in the study performed the randomization, pulled the assigned doses, checked the certificate of analysis, and over labeled with the randomization code. For randomization to VMT, the staff member also confirmed that the CFU count had been performed within the past month, otherwise, completed that testing to ensure viability prior to using the dose.

Participants returned within 48 hours of completing metronidazole for their first dose of study product. A second dose was delivered 36-72 hours after the first one. Participants were required to avoid sexual intercourse or insertion of any vaginal products for 72 hours prior to intervention visits. The study product was thawed for 30 minutes at room temperature prior to use. To deliver the intervention, a speculum was placed, and a transfer pipette with a wide aperture used to aspirate the fluid from the cryovial and place it in the vagina. The speculum was then turned sideways and angled to allow the fluid to drain out of the cup of the posterior blade and slowly removed. The participant then lay flat for 15 minutes.

Participants returned 1, 2 and 4 weeks after transplant, and then 2, 4 and 6 months later. Vaginal swabs were collected at the beginning of each visit. At V02 (pre-MTZ), V03 (post-MTZ), V05 (1 week post-VMT), V07 (1 month post-VMT) and V09 (4 months post-VMT), disposable menstrual disc was also inserted for 15-20 minutes to collect vaginal fluid. At the baseline (V02) and 4-week/1-month visit (V07), an endocervical cytobrush was collected and blood was drawn to measure immune cell populations. Participants were also tested for human immunodeficiency virus, herpes simplex virus, syphilis, Hepatitis B, Hepatitis C, Cytomegalovirus, *Neisseria gonorrhoea, Chlamydia trachomatis, Trichomonas vaginalis, Candida albicans,* and *Mycoplasma genitalium* at V02 and V07. At each visit, participants also completed questionnaires about symptoms, recent sexual activity, recent vaginal product use, and antibiotic use. Between visits, participants were asked to call if they had any new or worsening symptoms; these data were used to assess for adverse events.

### Menstrual disc processing

After collection, menstrual discs were placed into 50 mL conical tubes and centrifuged at 810 RCF for 10 minutes to collect vaginal fluid in the bottom of the tube. The menstrual discs were then discarded, and the samples were weighed by taring an analytical balance with an empty menstrual disc inside of a 50 mL conical tube. Each vaginal fluid sample was then diluted with 0.5 mL of sterile saline and sheared with a blunt-end 16-gauge needle on a syringe. The volume of each sample was estimated to the nearest 0.1 mL after being taken up into the syringe, and the samples were then aliquoted into cryovials and frozen at - 80°C.

### Cytokine measurement and analysis

Pre-processing - Vaginal fluid samples were thawed on wet ice, and then 100µLof each sample was diluted with 100µLof PBS. These dilutions were centrifuged at 1000 RCF for 15 minutes. 100µLof supernatant was removed from each sample and transferred to a Corning® FiltrEX 96-well Filter Plate that was then centrifuged at 2451 RCF for 1 hour. The flowthrough was frozen at -80°C.

The concentrations of 20 cytokines/chemokines (MIG, IP10, IFN-γ, ITAC, IL1α, IL1β, TNFα, IL6, IL8, MIP-3α, IL12, MIP1α, MIP3β, IL13, IL 12, IL21, IL4, IL23, IL5, IL10) were measured in the vaginal supernatant using multiplexed ELISA assays (Luminex), as previously described (*3*). Values below the lower limit of detection in the assay were imputed as half of the lowest standard concentration for that analyte. Similarly, concentrations above the detectable limit were imputed as twice of the highest standard concentration.

### Flow Cytometry

Flow cytometry was used to evaluate alterations in the endocervical immune cell populations with the change of vaginal microbiota. Vaginal cytobrush samples were processed within 2 hours after collection. Cells were stained with LIVE/DEAD fixable blue dead cell stain dye and fluorescent monoclonal antibodies specific for the following human surface markers: CD45 (Alexa700), CD8 (APC-H7), CD56 (BV711), CD3 (PE-CF594), CD66b (PE-A), MHCII (FITC), CD4 (BV786), CD1a (BV650), CD14 (BV605), CD19 (BV510), CD11c (BUV737), and CD16 (BUV395). Samples were then fixed with 1% paraformaldehyde (PFA) and analyzed on the flow cytometer 5-laser LSR Fortessa (5L Fortessa) (BD) within 48 hours. We used the following controls for each experiment: an unstained sample and calibration (“rainbow”) beads. Data analysis was conducted using FlowJo Version 7.0.0 (FlowJo Enterprise). Viability dye was used to exclude non-viable cells, singlet gating was used to exclude aggregates. Sequentially the cells were gating as follow: the granulocytes were defined as CD45+ CD66b+, the CD4 and the CD8 T cells CD45+, CD66b-,CD3+ and respectively CD4+ or CD8+, the B cells as CD45+, CD66b-,CD3-, CD19+, the NK+-cells as CD45+, CD66b-,CD3-, CD19-, CD56+, the Monocytes as CD45+, CD66b-,CD3-, CD19-, CD56-, MHCII +, CD14+, the Langerhans cells as CD45+, CD66b-,CD3-, CD19-, CD56-, MHCII+, CD14+, CD16-, CD123+, the plasmacytoid cells as CD45+, CD66b-,CD3-, CD19-, CD56-, MHCII+, CD14+, CD16-, CD123-, CD1a+, CD1c+, and the conventional DCs as CD45+, CD66b-,CD3-, CD19-, CD56-, MHCII+, CD14+, CD16-, CD123-, CD1a-, CD1c+, MHCII high. All populations are expressed as percentage of CD45+ cells.

### Secretor status

The human secretor status is controlled by the FUT2 gene. The G428A (W-stop codon) mutation on this gene is the most frequent allele responsible for non-secretor status in Caucasian and in Black populations. The secretor status was identified using a method develop by Marionneau et al. (*19*). It takes advantage of the G428A mutation overlapping the AvaII restriction enzyme binding site. The total nucleic acid extracted from 25ul inoculum aliquots (see method described below) contained a significant amount of DNA from vaginal epithelial cells. A fragment of the FUT2 gene, containing the position 428, was amplified with the primers 5’—CCATGCTGGTCGTTCAGATGCCTTTCTCCT-3’ and 5’-GCTCATGGAACCATGTGCTTCTCATGCCCG-3’ as described by Soejima et al. (*20*) and was digested with Ava II. The G428A mutation abrogates one of the restriction sites of this enzyme The change of the AvaII restriction pattern was visualized on agarose gel.

### DNA extraction from swabs

Swabs were eluted in 800µL sterile saline and vortexed thoroughly. An aliquot of 200µL was used for DNA extraction. Total nucleic acids was extracted using a phenol-chloroform method adapted for 96-well plates, and included bead beating to disrupt bacterial cell walls (*21*). Genomic DNA from culture isolates was extracted with a plate-based protocol combining bead beating, phenol-chloroform isolation, and the QIAamp 96 DNA QIAcube HT kit (*22*).

### Microbiome profiling using 16S rRNA amplicon sequencing

The V4 region of the bacterial 16S rRNA gene was amplified with primers 515F/806R (with 806R barcoded for multiplexing) and sequenced on an Illumina MiSeq with a 300-cycle kit (*3, 21, 22*). Taxonomic assignment was performed with dada2 using the RDP database v16.

### Quantitative PCR

*L. crispatus* was quantified using the TaqMan Gene Expression Assay ID Ba04646245_s1 (ThermoFisher), with a calibration curve generated from whole genomic DNA. *L. iners* was quantified with a previously validated TaqMan assay and plasmid standard curve (*23*). Total bacterial abundance was measured with the BactiQuant qPCR as described previously (*24*).

### Bacterial isolations

Frozen vaginal microbiota donations were thawed on wet ice. Ten microliters of donation fluid was resuspended in 90 µL of sterile PBS, serially diluted (10⁻³ - 10⁻⁶), and drop-spotted (3 × 10 µL) onto de Man, Rogosa, and Sharpe (MRS) agar plates. Plates were incubated anaerobically at 37°C for 48 hours, and colonies from the most countable dilution (<30 per drop) were enumerated and imaged.

Ten colonies per sample from the 10⁻⁵ and 10⁻⁶ dilutions were. randomly selected, sub-cultured on half MRS plates, and incubated anaerobically for 72 hours. Isolates were re-streaked once more on full MRS plates to ensure purity. Single colonies were inoculated into MRS broth, and used to generate glycerol stocks and 1 mL pellets for downstream DNA extraction and whole-genome sequencing. DNA extractions were optimized for long-read compatibility.

### Bacterial genome sequencing

Culturing and pelleting: *L. crispatus* isolates from glycerol stocks were grown anaerobically on MRS agar (Hardy Diagnostics #G117) at 37°C for 48 hours. Colonies were inoculated into 3mL MRS broth (BD DifcoTM #288130) and incubated anaerobically overnight (∼18 hours) at 37°C. One-milliliter aliquots were pelleted (5000 x g, 5 minutes, 4°C), supernatant removed, and pellets stored at -20°C.

DNA extraction and sequencing (long read): Pellets were resuspended in 5M lithium chloride (Molecular Dimensions #MD2-100-43), washed with PBS, and processed with the MasterPure^TM^ Gram Positive DNA Purification Kit (Biosearch Technologies #MGP04100). DNA was eluted into 35µL TE buffer, quantified with a Qubit^TM^ 4 Fluorometer (Invitrogen #Q33226), diluted to 200 ng/10𝜇L, and libraries prepared with Oxford Nanopore’s Rapid Barcoding Kit (SQK-RBK114.24). Barcoded DNA was pooled, purified, and loaded onto MinION flow cells using R10.4.1 chemistry (FLO-MIN114) using a MinION Sequencing Device (MIN-101B). Sequencing ran for 48-72 hours, generating 3.5-11Gb of bases.

Assembly and polishing: Reads were demultiplexed (MinKNOW v24.02.8), concatenated if from multiple runs, and filtered for quality and length using Filtlong v0.2.1. Filtered reads were subsampled into 12 sets with Trycycler v0.5.3, and deduplicated with BBMap v39.00. Assemblies were generated using Flye v2.9.1, Minipolish v0.1.3, Raven v1.8.1, and Canu v2.2, then clustered and reconciled with Trycycler. Manually selected clusters were curated based on read depth, number of contigs, and sequence identity of assemblies as described in the Trycycler documentation (*25*). A consensus contig sequence for each cluster was generated based on a multiple sequence alignment and read partitioning using Trycycler. Consensus contigs were polished with Medaka v1.7.2 (long reads) and Polypolish v0.5.0 (short reads). Assembly quality was assessed with Busco v5.5.0.

### Metagenomic sequencing and culture isolate whole-genome sequencing

Libraries were prepared with the Nextera DNA Library Preparation Kit (Illumina) and KAPA HiFi Library Amplification Kit using a modified protocol (*26*). DNA was quantified with SYBR Green, normalized to 1ng/1 µL, and then simultaneously fragmentation and tagmented by mixing the DNA with 1.25µLTD buffer and 0.25µLTD Enzyme 1 and incubating for 9 minutes at 55°C. Tagmented fragments were PCR-amplified with KAPA reagents and Illumina adaptor sequences and sample barcodes incorporated in the primers. PCR products were pooled, bead-purified and sequenced on an Illumina NovaSeq X (300-cycle kit).

### Bacterial genomics

Isolate genome assemblies were used to construct whole-genome phylogenies with Parsnp (Harvest suite) (*27*), with the recombination filtering (-x). Alignments (.ggr format) were converted to a SNP FASTA alignment using HarvestTools. Maximum-likelihood phylogenies were inferred with IQ-TREE (*28*) under default model selection and optimization.

### Metagenomic analyses

Shotgun libraries were sequenced on an Illumina NovaSeq. Cutadapt version 4.8 was used to remove adapters. Trimmed reads were quality controlled using sickle version 1.33 with a minimum quality of 20 and minimum read length of 50 bps. Human reads were removed using a two-step filtering process against the T2T human reference, where first bbduk version 39.01, a kmer-based read filtering tool, was used and then reads were further filtered using HISAT2 version 2.2.1, an alignment-based mapper.

Taxonomic profiling was performed with VIRGO2 (*18*). Reads were mapped to the VIRGO2 bowtie database and reads unambiguously mapping to species-specific genes were used to estimate the abundance of the species, taking into account the length of each species-specific gene which recruited reads. To compare gene content between donations and participant samples, genes annotated as specific to *L. crispatus* in VIRGO2 with coverage > 10% the median coverage were considered present in a sample. For isolate-metagenome comparisons (purple heatmap in Figure 3), reads from metagenomic samples were mapped to isolate genomes using bbmap version 39.01.

