## Supplemental Material for "Donation strain engraftment demonstrates feasibility of vaginal microbiota transplantation to prevent recurrent bacterial vaginosis"

**Supplemental Table S1. Adverse events for pilot study.**

| | VMT | Placebo |
|---|---|---|
| **Serious adverse events** | 0 | 0 |
| **Adverse events grade 2 or higher** | 2 | 2 |
| **Adverse genitourinary events grade 2 or higher** | 0 | 2 |
| **Urogenital events of any grade** | | |
| Urinary frequency | 1 | 0 |
| Vaginal odor | 1 | 1 |
| Painful sex | 1 | 0 |
| Vaginal discharge | 1 | 1 |
| Vulvovaginal irritation | 1 | 0 |
| Yeast infection | 0 | 1 |
| Vaginal spotting | 0 | 1 |

**Fig. S1**. **Donation weight and volume measurements**

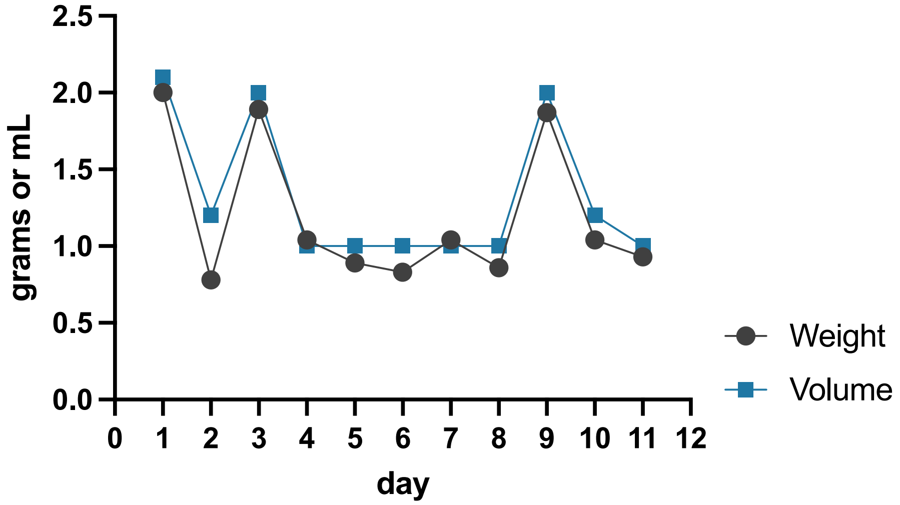


**Fig. S1**. **Donation weight and volume measurements.** Volume (mL) plotted in blue, and weight (g) plotted in grey of donation fluid for each total donation (one donation each day)**.**

**Fig. S2. Recipient microbiome community dynamics using 16S rRNA sequencing.**

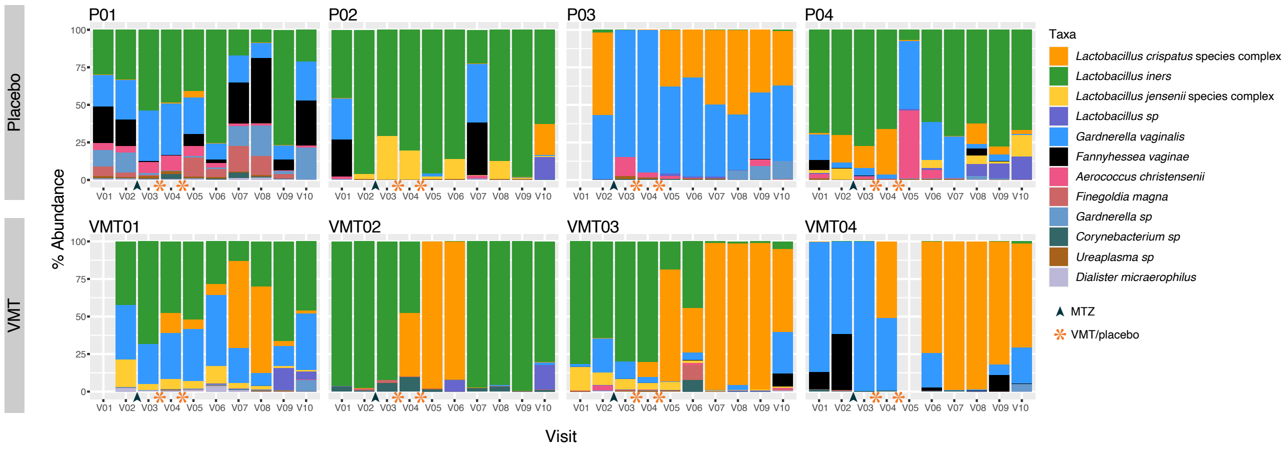


**Fig. S2. Recipient microbiome community dynamics using 16S rRNA sequencing.** Relative abundance of microbial taxa in Placebo recipients (above) and VMT recipients (below) calculated using 16S rRNA gene amplicon sequencing. Metronidazole administration indicated with black arrow and treatment indicated with orange asterisk. Primary outcome for trial determined at 1-month post-treatment (V07).

**Fig. S3**. **Relative abundance of *L. crispatus* alone.**

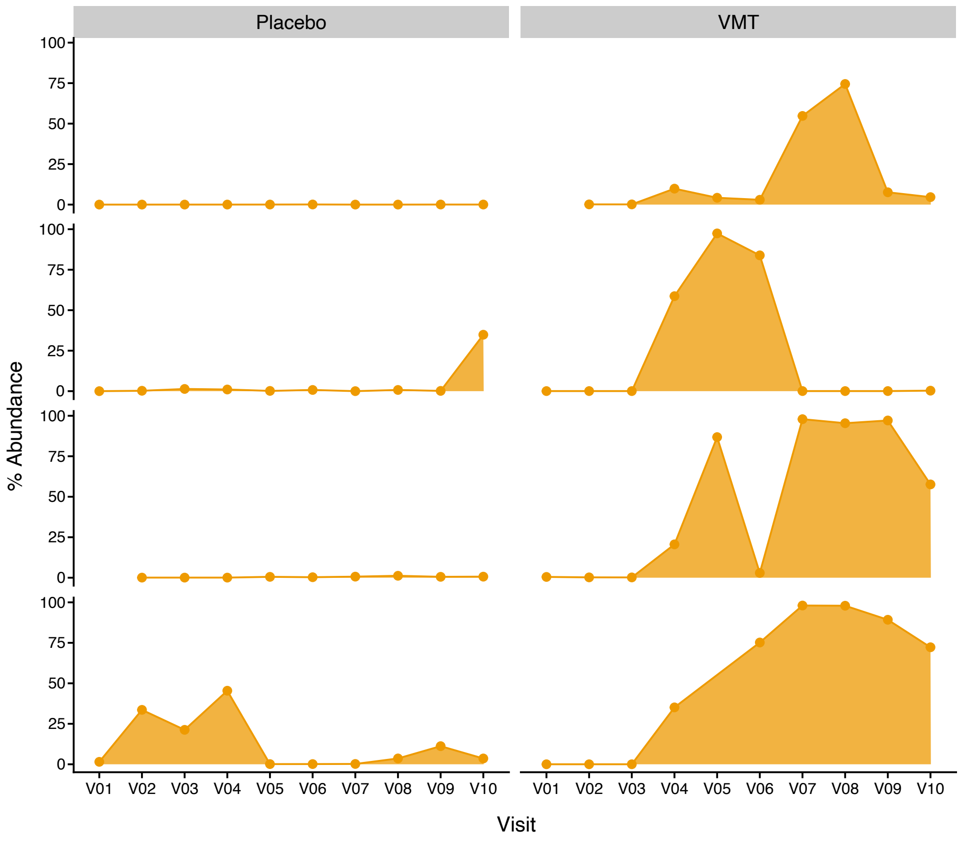


**Fig. S3**. **Relative abundance of *L. crispatus* alone.** Metagenomically-determined relative abundance of *L. crispatus* in each sample (taxonomic assignments from VIRGO2).

**Fig. S4**. **Absolute abundance of select taxa and cytokines.**

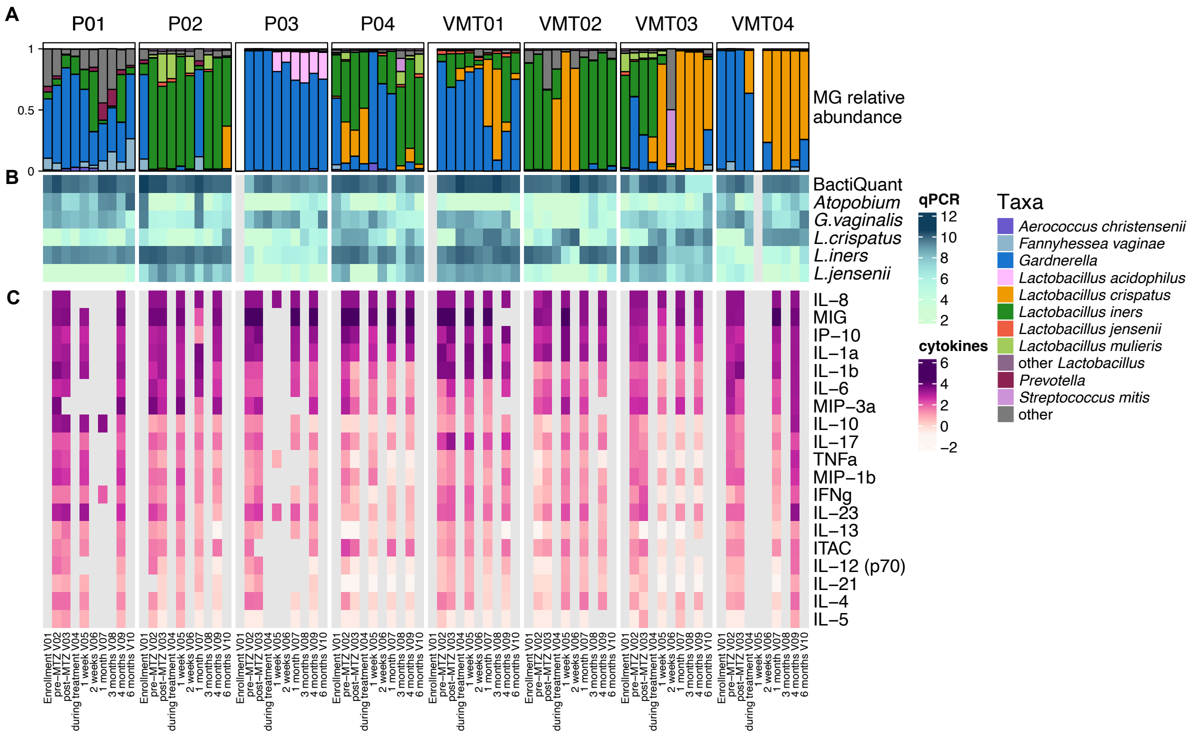


**Fig. S4**. **Absolute abundance of select taxa and cytokines.** (A) Relative abundance of each sample determined from metagenomics using VIRGO2 as a reference database. (B) Absolute abundance measured using species-specific qPCR assays in copies/swab. (C) Total cytokine concentration (pg/uL) in each sample assay.

**Fig. S5**. **Change from baseline of cytokines**

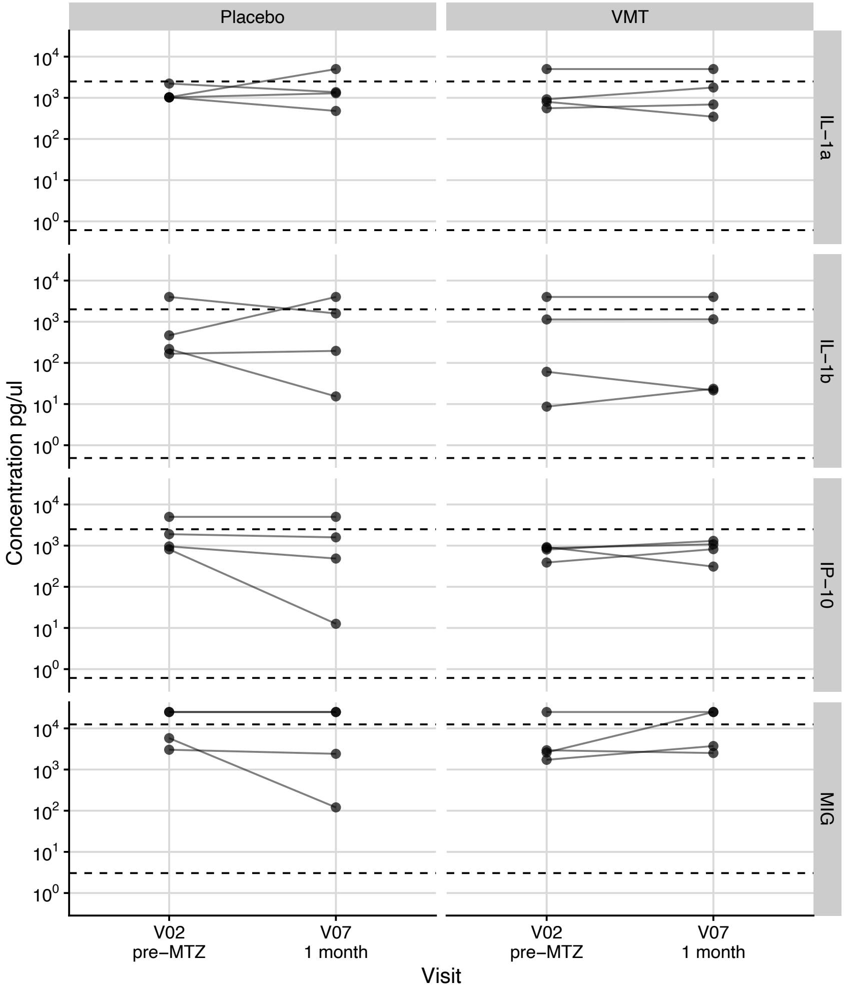


**Fig. S5**. **Change from baseline of cytokines.** Select cytokines (right) compared between baseline (V02) and 1-month post-treatment (V07).

**Fig. S6**. **Immune cell concentrations.**

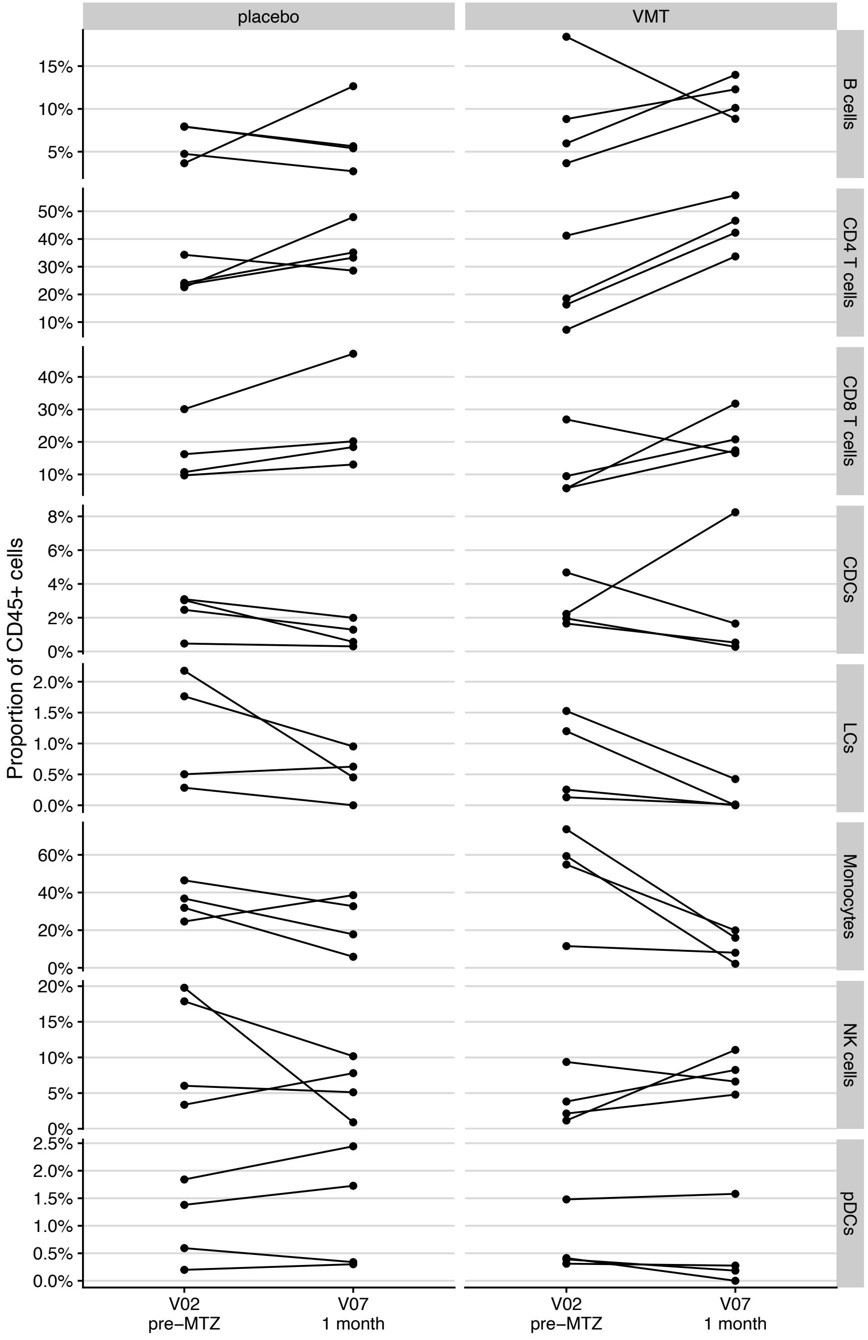


**Fig. S6**. **Immune cell concentrations.** Select immune cells (right) compared between baseline (V02) and 1-month post-treatment (V07) as proportion of CD45+ cells.
